# Association Between Depression and All-Cause Mortality : A Cohort Study Based on NHANES Data

**DOI:** 10.1101/2025.04.29.25326650

**Authors:** Fang Yao, Jiayao Zhang, Fang Yao, Hui Wang

## Abstract

**Background:** Depression is a common mental disorder that significantly influences mental health outcomes and increases the risk of all-cause mortality. However, the dose-response relationship between depression and all-cause mortality remains unclear.

**Methods:** A total of 36,193 participants from the National Health and Nutrition Examination Survey (NHANES) 2005–2018 were included in this study. Depressive symptoms were assessed using the Patient Health Questionnaire-9 (PHQ-9) scale, and all-cause mortality data were obtained from the National Death Index. Weighted Cox regression models were applied to evaluate the association between depression and all-cause mortality, with special attention to non-linear relationships.

**Results:** Among the study population, 8.74% were identified as having depression. The mean follow-up duration was 90.53 months, during which 9.87% of participants died from all causes. A significant non-linear association was observed between PHQ-9 scores and all-cause mortality. All-cause mortality increased markedly with depression scores ≤ 7 (HR: 1.068, 95% CI: 1.042–1.095, *P* < 0.0001) and plateaued when scores approached 7.

**Conclusions:** Elevated depressive symptoms, even at mild levels, are associated with a substantial increase in all-cause mortality. These findings underline the necessity for early identification and intervention for depressive symptoms to reduce long-term adverse health outcomes. This research provides critical evidence to inform public health strategies targeting the reduction of depression-related mortality.

## Introduction

Depression is acknowledged as a prevalent mental health disorder affecting over 300 million people globally. It is increasingly recognized as a key contributor to premature mortality[1]. The World Health Organization emphasizes that the ramifications of depression extend beyond mental health, significantly impacting physical health and longevity. Epidemiological studies consistently associate depression with heightened risks of mortality from various causes, including cardiovascular diseases, cancer, and suicides. For instance, a comprehensive meta-analysis involving 293 cohort studies has revealed that individuals diagnosed with depression have a 50% increased risk of all-cause mortality, with even greater ascertainments noted for cardiovascular-related deaths[2,3]. This relationship underscores the urgent need for public health strategies aimed at early identification and management of depression, particularly given its profound implications for overall mortality.

Despite these alarming statistics, the mechanisms that link depression to increased mortality remain somewhat elusive. Research suggests that dysregulated neuroendocrine pathways, chronic inflammation, and alterations in health behaviors may underpin this association [4]. Notably, inflammation has emerged as a significant factor, potentially mediating the effects of depression on mortality [5]. Recent studies have confirmed that markers of systemic inflammation, such as elevated C-reactive protein levels, correlate with depressive symptoms and present associated mortality risks [6,7]. For example, Gialluisi et al. have highlighted how depression influences hospitalization and mortality risk through inflammatory mechanisms, emphasizing that systemic inflammation accounts for a notable proportion of the risk associated with depression [8]. Nonetheless, research often predominates from clinical or elderly populations, limiting the extrapolation of these findings to community-dwelling adults who equally experience depression and its detrimental effects on health [9].

To address these research gaps, the National Health and Nutrition Examination Survey (NHANES) presents a unique opportunity to investigate the association between depression and mortality across diverse sociodemographic groups. NHANES incorporates nationally representative sampling, long-term mortality follow-up, and comprehensive biomarkers, allowing for a multifaceted analysis of how depression affects mortality rates [10].Prior NHANES-based research has discerned significant links between depressive symptoms and conditions such as metabolic syndrome, systemic inflammation, and heightened levels of oxidative stress, all of which play vital roles in mortality pathways [11,12] . However, despite these insights, much of the existing literature has focused on cross-sectional relationships rather than long-term mortality risks, which poses limitations on understanding the full implications of depression over time.

This study aims to elucidate the longitudinal relationship between depression and all-cause mortality in the NHANES population, highlighting the urgency of integrating mental health interventions into public health frameworks to reduce the elevated mortality rates among individuals with depression.

## Materials and methods

### Study population

The National Health and Nutrition Examination Survey (NHANES), a national research program conducted by the National Center for Health Statistics (NCHS) of the Centers for Disease Control and Prevention (CDC), is designed to assess the health and nutritional status of the civilian, non-institutionalized U.S. population. Using a stratified multistage sampling design, NHANES collects data on approximately 5,000 persons annually through household interviews and standardized physical examinations in mobile examination centers (MECs). The survey, which includes demographic, socioeconomic, dietary, and health-related data, is released in 2-year cycles. Written informed consent is obtained from all participants, ensuring ethical compliance. This methodology allows NHANES to provide accurate estimates of disease prevalence and effectively represent the U.S. population [13–16].

This study used survey data from 2005 to 2018. Participants were excluded from analyses if they were <18 years old and had incomplete data on depression and all-cause mortality.

### Depression assessment

Depression was measured in NHANES using the Patient Health Questionnaire-9 (PHQ-9), a 9-item screening tool designed to assess depressive symptoms experienced over the past two weeks. Each item is scored from 0 to 3 points, which correspond to the frequency of symptoms experienced, with total scores ranging from 0 to 27 points. Higher scores indicate more severe depressive symptoms. The PHQ-9 serves as both a screening and diagnostic tool, demonstrating robust psychometric properties, with studies confirming its effectiveness in various populations [17,18]. Consistent with previous studies, a PHQ-9 total score of ≥10 points is deemed clinically relevant, indicating significant depressive symptoms and aligning with established diagnostic thresholds in mental health [19].

### All-cause mortality assessment

The present study utilized the public use of the Linked Mortality File (LMF) from the National Death Index (NDI) as of 31 December 2019 to ascertain mortality data. Mortality data for NHANES participants aged 20 years and older were acquired through 31 December 2019, from the NHANES Public-Use Linked Mortality File, which is linked to the National Death Index (NDI) via a probabilistic algorithm. The causes of death were coded in accordance with the International Classification of Diseases, 10th Revision (ICD-10), and the study endpoint was defined as all-cause mortality. The NHANES 2005-2018 Linked Mortality Files (LMFs) were correlated with the 2019 National Death Index death certificate records by deterministic and probabilistic methods, and follow-up was continued until death. Participants who were not matched to a death record were considered to be alive. Relevant data are available through the NCHS Research Data Centre [20].

### Covariate assessment

In order to assess the impact of potential confounders, a selection of important covariates was made, including sex, age, race, family poverty income ratio (PIR) (<1.30, 1.30-3.50, ≥3.50) [21], education levels (less than 9th grade/9-11th grade (includes 12th grade with no diploma)/high school graduate/some college or AA degree/college graduate or above), and marital status (married/living with partner/never married/divorced/separated/widowed), body mass index (BMI) (<25, 25-30, ≥30) [22].These covariates were collected using standardised questionnaires. Each participant’s weight and height were determined by physical examination.Body mass index (BMI)was defined as weight (kg) divided by height squared (m^2^). Among 36,193 patients, the number of missing values for covariates was as follows: Education Levels 2,155 (5.95%), Marital Status 1,634 (4.51%), Family PIR 3,061 (8.46%), and BMI 384 (1.06%).

### Statistical analysis

All analyses were conducted utilising the statistical software packages R (http://www.R-project.org, The R Foundation) and EmpowerStats (http://www.empowerstats.com, X&Y Solutions, Inc, Boston, MA). It is imperative to note that all tests were two-sided, and *P* values lower than 0.05 were considered statistically significant.

The baseline characteristics of all patients were grouped according to the presence or absence of depression.Continuous variables were analysed using a weighted linear regression model, and categorical variables were analysed using the Chi-square test to compare differences between the depressed and non-depressed groups.The initial characteristics of the subjects were described in accordance with their depressive status. Mean and standard error (SE) were employed to describe continuous variables, while numbers and percentages were used to describe categorical variables. Kaplan-Meier survival curves and log-rank tests were used to visualize and compare survival probabilities across PHQ-9 score groups. The cox model with restricted cubic spline demonstrated a dose-response relationship between depression scores and the risk of death. A two-stage modelling approach was used for the primary analyses: standard linear regression (model 1) and two-part linear regression (model 2). Hazard ratios (HRs) with 95% confidence intervals (CI) were calculated to quantify the risk of death after adjustment for survey weights and potential confounders. The primary outcome measure was the adjusted HR for each 1-point increase in PHQ-9 score. Secondary analyses included stratified subgroup analyses by demographic and clinical characteristics to explore effect modification. Model performance was assessed using the log-likelihood ratio test, and statistical significance was defined as *P* < 0.05.

In order to ascertain the robustness of the results, sensitivity analyses were conducted. The utilisation of dummy variables was necessitated by the absence of continuous variables by more than 1% [23].

## Result

### Participant characteristics

The present study analysed data from the NHANES surveys conducted between 2005 and 2018. From the initial cohort of 101,318 participants, 28,047 were excluded on the basis that they were < 18 years, leaving 73,271 potential participants. Subsequently, 37,012 participants were removed due to missing depression data, and an additional 66 were excluded for incomplete all-cause mortality information. The final analytical sample comprised 36,193 participants (Fig 1).

**Fig 1.**
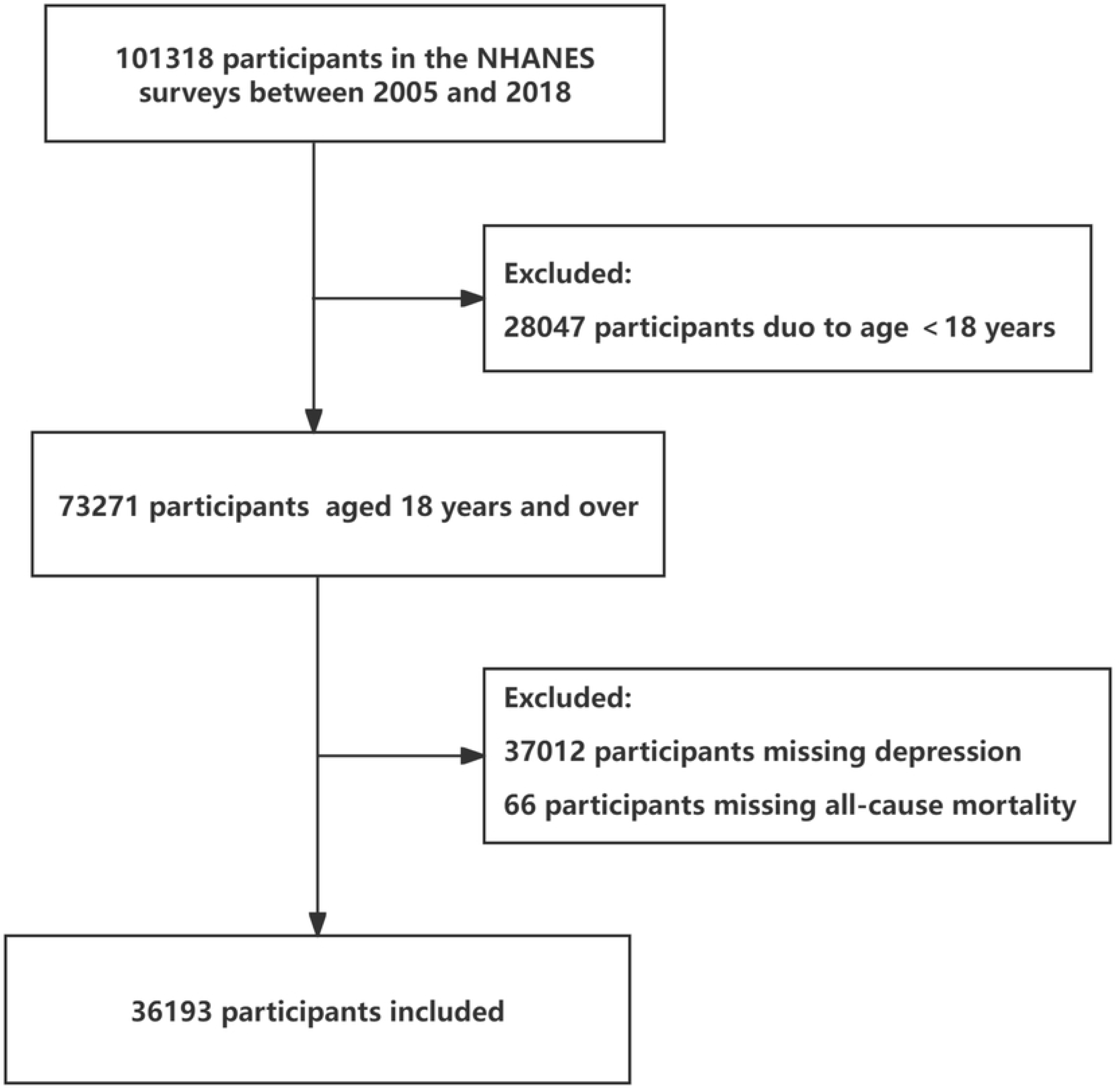
Flow chart of study participants.

The findings of this study demonstrated significant disparities in baseline characteristics between the depression group and the non-depression group(Table 1). The depression group exhibited a higher mortality rate (12.39% vs. 9.63%, *P* < 0.001), significantly lower household income levels (*P* < 0.0001), and a higher BMI (*P* < 0.0001). Additionally, the mean follow-up time was significantly shorter in the depression group compared to the non-depression group (P < 0.0001). Furthermore, a substantial disparity was observed in demographic characteristics, including gender distribution, educational attainment, and marital status (*P* < 0.001). Specifically, the depression symptoms group had a higher proportion of females (63.24% vs. 49.68%, *P* < 0.001) and lower educational attainment, with fewer individuals holding a college graduate degree or higher (10.086% vs. 24.130%, *P* < 0.001). Furthermore, the depression group exhibited significantly higher proportions of individuals who were single, divorced, or separated compared to the non-depression group.

**Table 1.**
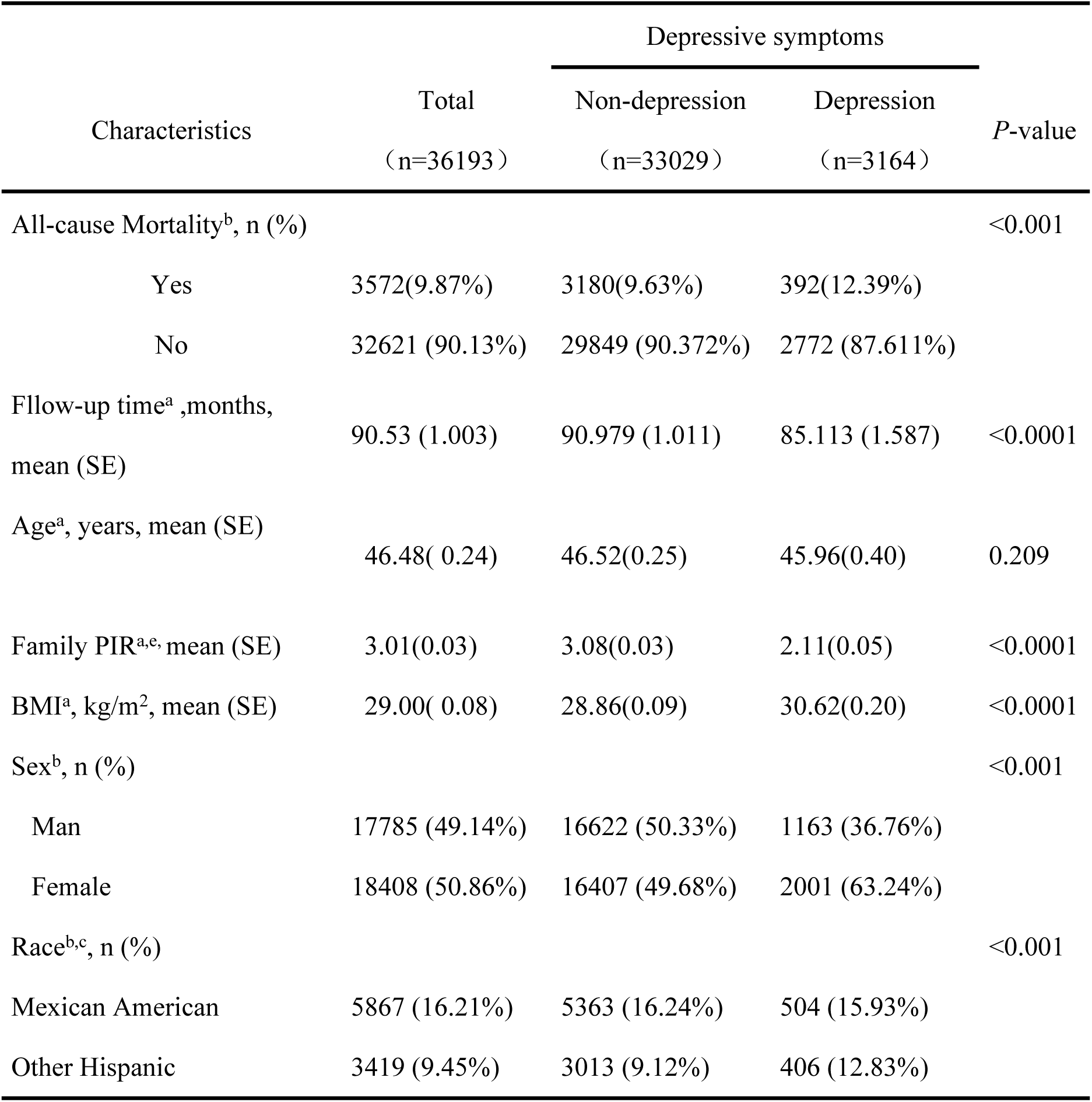
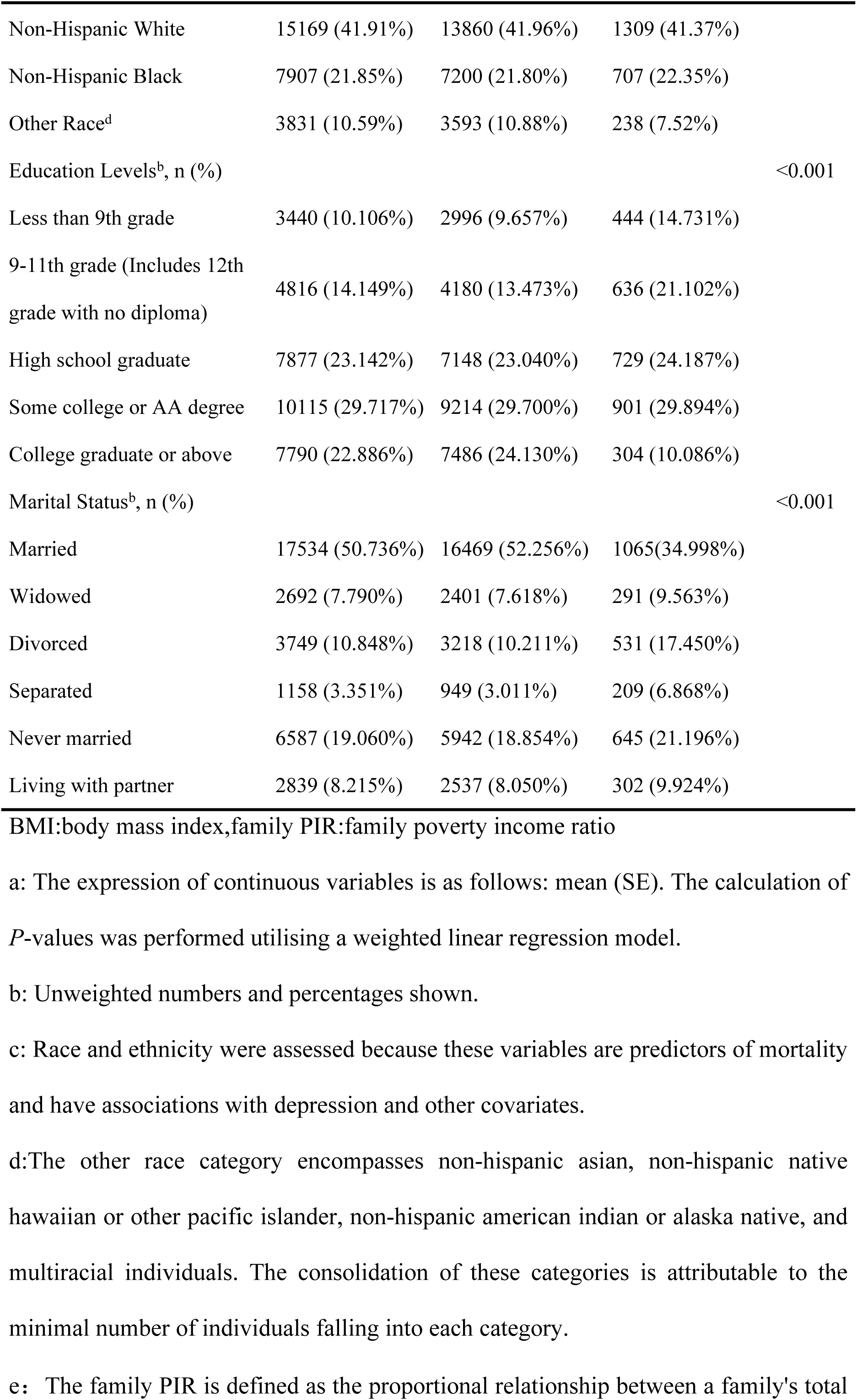
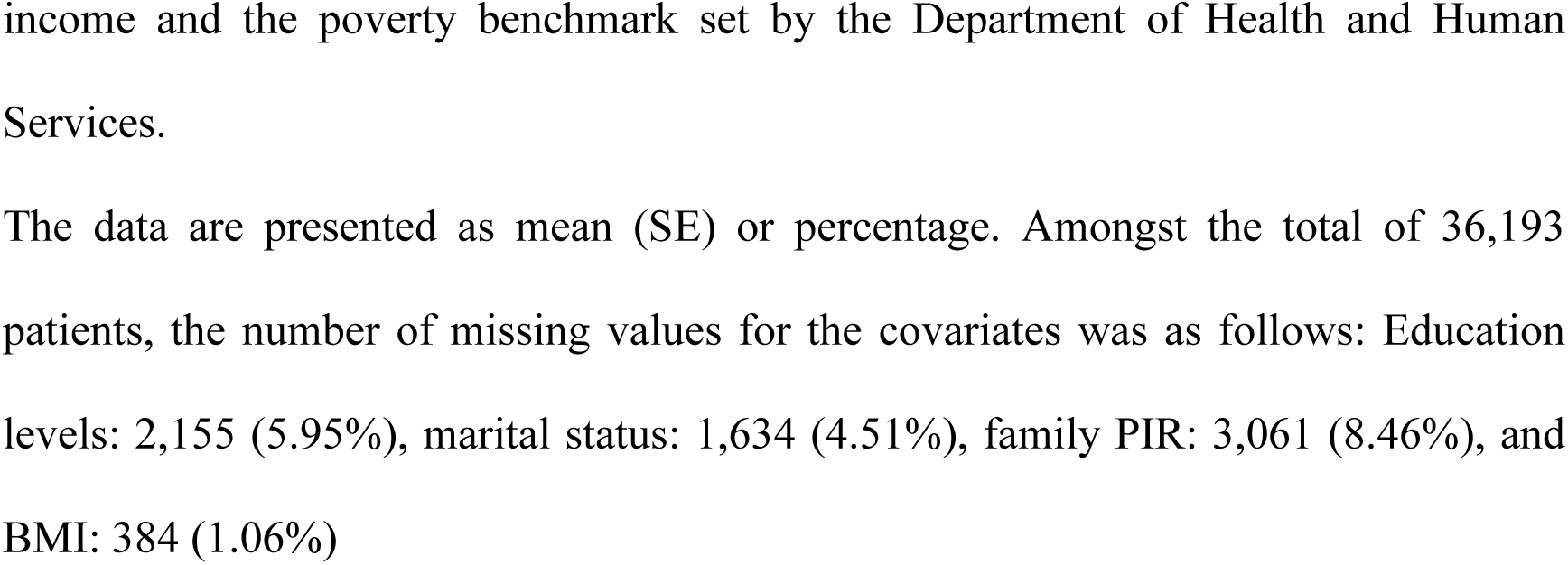
Baseline Characteristics of Study Participants Stratified by Depression Status ,Weighted.

The Kaplan–Meier curve indicated that patients diagnosed with depression were associated with an increased risk of all-cause mortality (*P* < 0.0001) (Fig 2).

**Fig 2.**
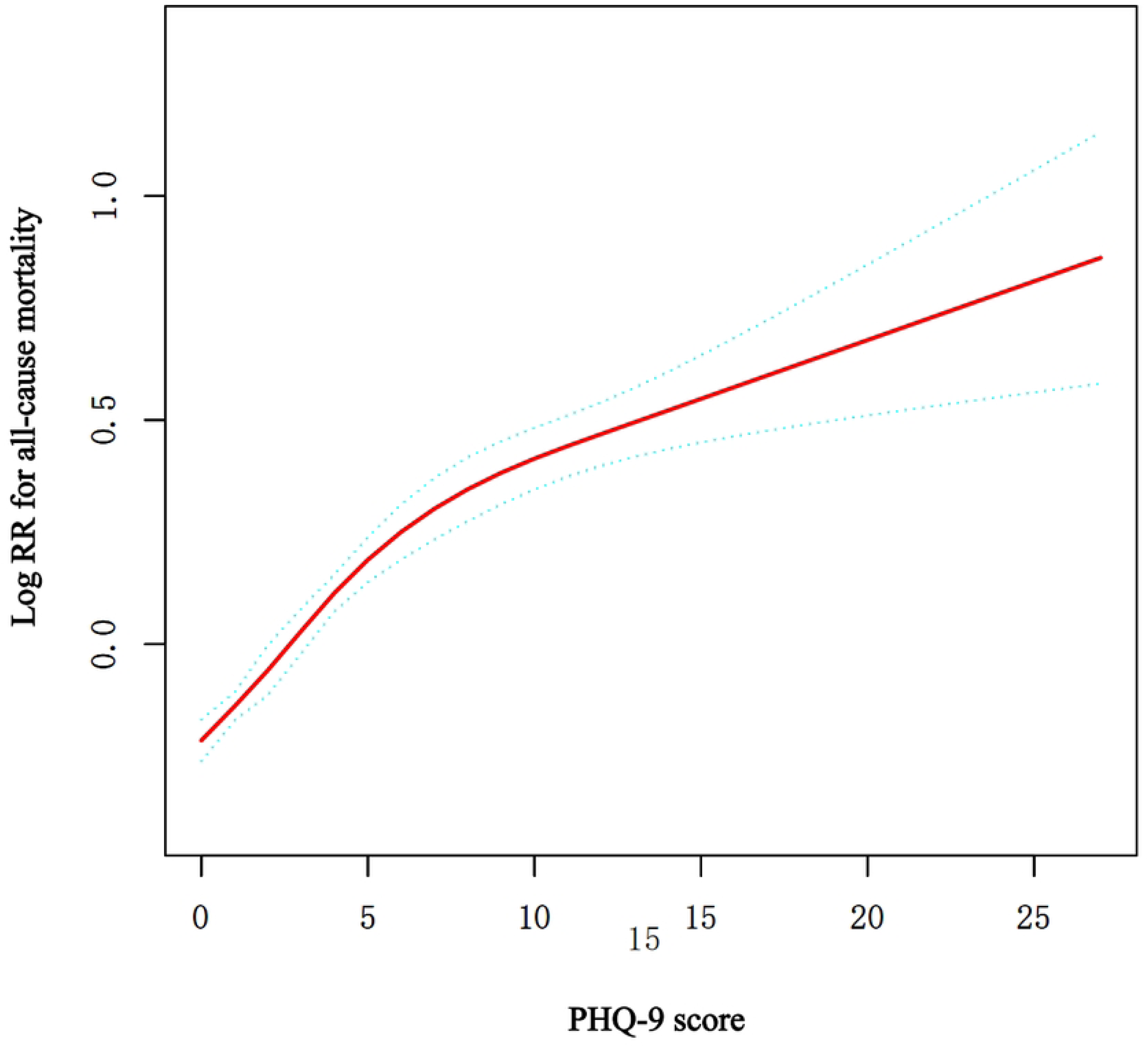
The Kaplan-Meier curves by depression status.

Fig 3 presented illustrate a non-linear relationship between depression, as measured by the PHQ-9 score, and all-cause mortality risk. The red line indicated the estimated Log RR for death, which showed a gradual increase as the PHQ-9 score rises. This suggested that higher levels of depression were associated with an increased risk of all-cause mortality. Importantly, this relationship remained significant after adjusting for potential confounding variables, including age, sex, BMI, race, marital status, family PIR, and education level. This adjustment strengthened the validity of the findings, suggesting that the observed association between depression and all-cause mortality was robust and not merely a result of these confounding factors.

**Fig 3.**
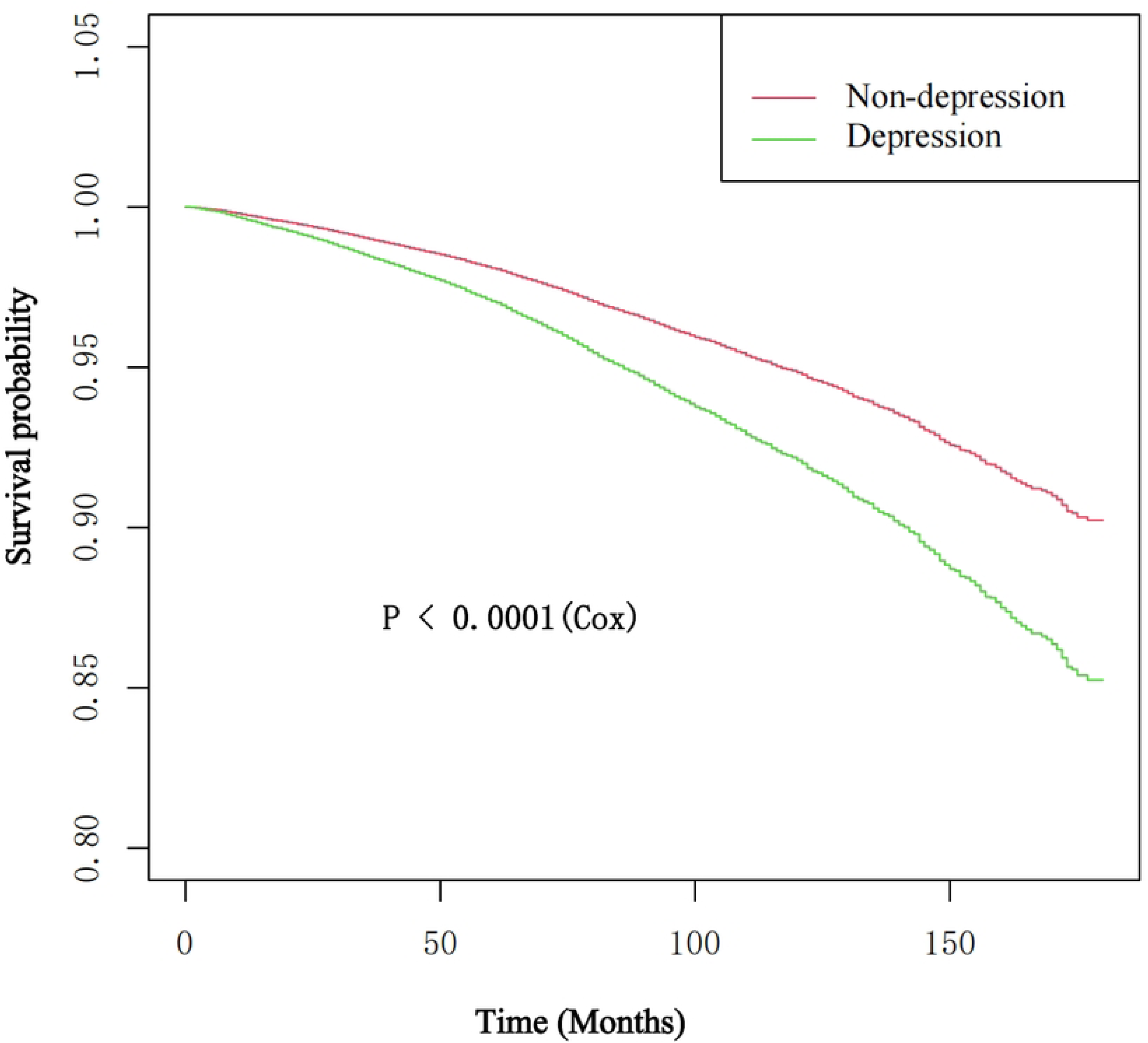
Association between depression and all-cause mortality.

Table 2 demonstrated a threshold effect of depression, as measured by the PHQ-9 scale, on the risk of all-cause mortality. Using a standard linear regression model (Model 1), a significant positive association was observed, with a 4.4% increased risk of all-cause mortality for each unit increase in PHQ-9 score (Adjusted HR: 1.044, 95% CI: 1.036–1.052, *P* < 0.0001). However, the two-piecewise linear regression model (Model 2) revealed a non-linear relationship, identifying a critical threshold at a PHQ-9 score of 7. For PHQ-9 score ≤ 7, the risk of all-cause mortality was significantly higher (Adjusted HR: 1.068, 95%CI: 1.042–1.095, *P* < 0.0001), while for PHQ-9 score >7, the risk increase was more moderate but still significant (Adjusted HR: 1.036, 95%CI: 1.008–1.064, *P* = 0.0113). The log-likelihood ratio test confirmed the superiority of the two-piecewise model (*P* < 0.001), suggesting that the association between depression and all-cause mortality risk was more pronounced at lower levels of depression. All models were adjusted for sex, age, race, family PIR, education levels, marital status, and BMI.

**Table 2.** Threshold effect analysis of depression on the risk of all-cause mortality using two-piecewise regression models.Weighted.

| Outcome | Adjusted HR(95%CI) | P-value |
| --- | --- | --- |
| Model 1 | 1.044 (1.036, 1.052) | <0.0001 |
| Model 2 |  |  |
| Inflection point of PHQ-9 score |  | 7 |
| ≤7 | 1.068 (1.042, 1.095) | <0.0001 |
| > 7 | 1.036 (1.008, 1.064) | 0.0113 |
| P for log-likelihood ratio test | <0.001 |  |

CI: confidence intervals, HR: risk ratios.
Threshold effect analyses for depression and all-cause mortality. The data were presented as HR (95%CI) *P*-values;
Model 1 was the linear analysis; model 2 was the nonlinear analysis. Adjustments had been made for factors including sex, age, race, family PIR, education levels, marital status, and BMI.
The findings revealed that Model 2 was significantly different from Model 1, with a *P* < 0.05.

Table 3 demonstrated a significant association between depression (measured by PHQ-9 score) and all-cause mortality, which was consistent across different adjustment models. As a continuous variable, each 1-point increase in PHQ-9 score was associated with a 2.6% higher risk of all-cause mortality in model 1 ( unadjusted) (HR:1.026, 95% CI: 1.019–1.033), 6.1% in model 2 (adjusted for sex, age, and race) (HR: 1.061, 95% CI: 1.054–1.069), and 4.4% in model 3(adjusted for sex, age, race ,family PIR, education levels, marital status, BMI) (HR: 1.044, 95% CI: 1.036–1.052), all with *P* < 0.00001. For PHQ-9 scores ≥10 (moderate-to-severe depression) compared to <10, the fully adjusted model demonstrated a significantly higher risk of all-cause mortality (HR: 1.547, 95% CI: 1.377–1.738, *P* < 0.00001). These findings indicated a dose-response relationship between depression severity and all-cause mortality risk, highlighting its independent and significant impact.

**Table 3.**
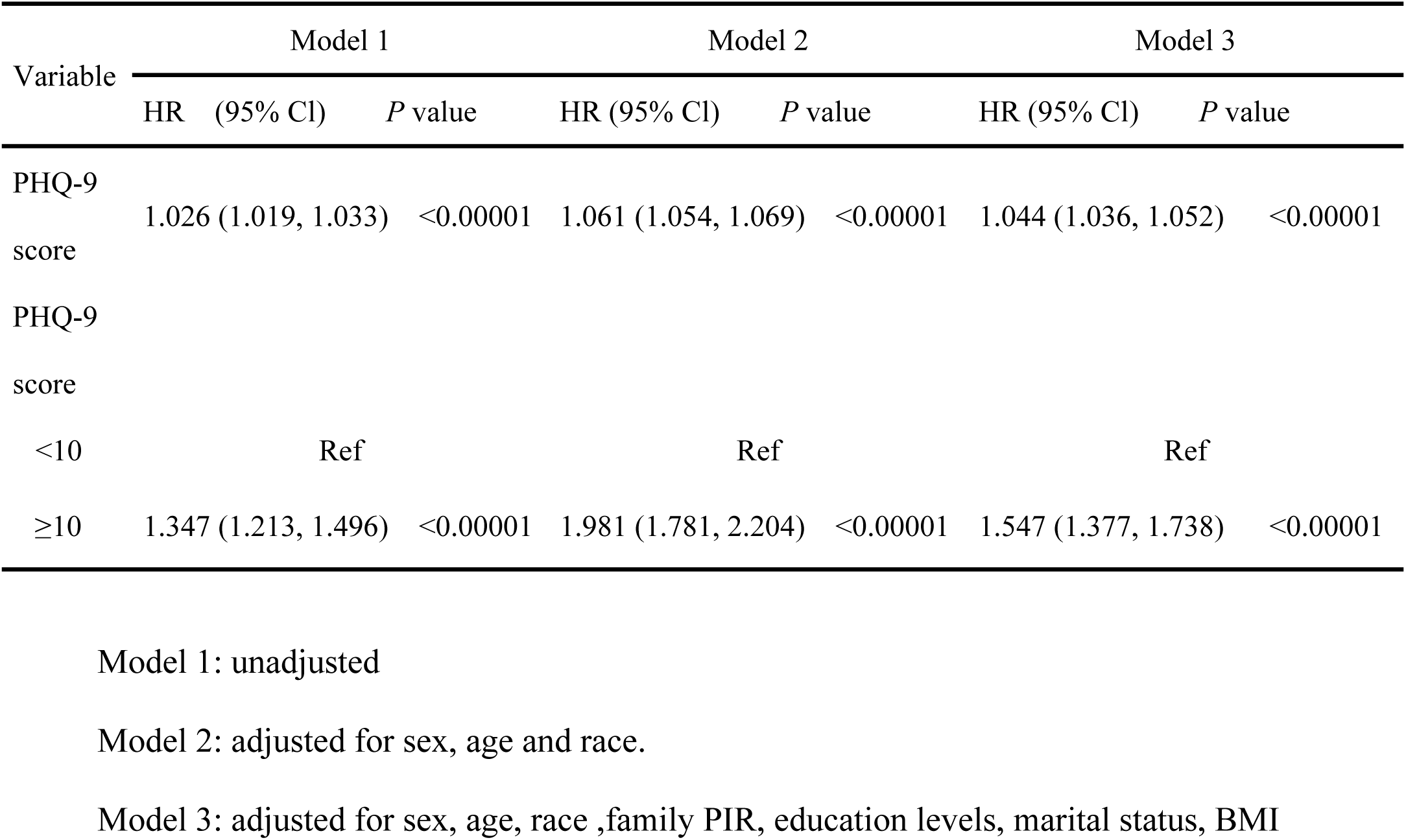
Relationship between depression and all-cause mortality in different models.Weighted.

The forest plot (Fig 3) illustrated the association between depression and all-cause mortality across different subgroups.After adjusting for sex, age, family PIR, BMI, education level, marital status, and race, the association between depression and all-cause mortality was significant across most groups (HR > 1, *P* < 0.05), with some stratification variables showing notable interaction effects. Specifically, age (*P*-interaction < 0.0001) and BMI (*P*-interaction = 0.004) demonstrated significant modification effects. Younger individuals (≤60 years) (HR: 1.064, 95% CI: 1.050–1.077) had a higher risk compared to older individuals (> 60 years) (HR: 1.027,95% CI: 1.017–1.037). Similarly, individuals with a BMI <25 (HR:1.062, 95% CI: 1.048–1.077) were at greater risk compared to those with a BMI of 25–30 (HR: 1.028, 95%CI: 1.013–1.042). Race also significantly modified the association (*P*-interaction = 0.005), with other race (HR: 1.057, 95% CI: 1.018–1.097) showing the highest risk. In contrast, sex (*P*-interaction = 0.7331), family PIR (*P*-interaction = 0.2958), education level (*P*-interaction = 0.2687), and marital status (*P*-interaction = 0.2002) had no significant interaction effects, indicating similar risks across these groups. Overall, depression significantly increased the risk of all-cause mortality, with age, BMI, and race being critical modifiers.

**Fig 3.**
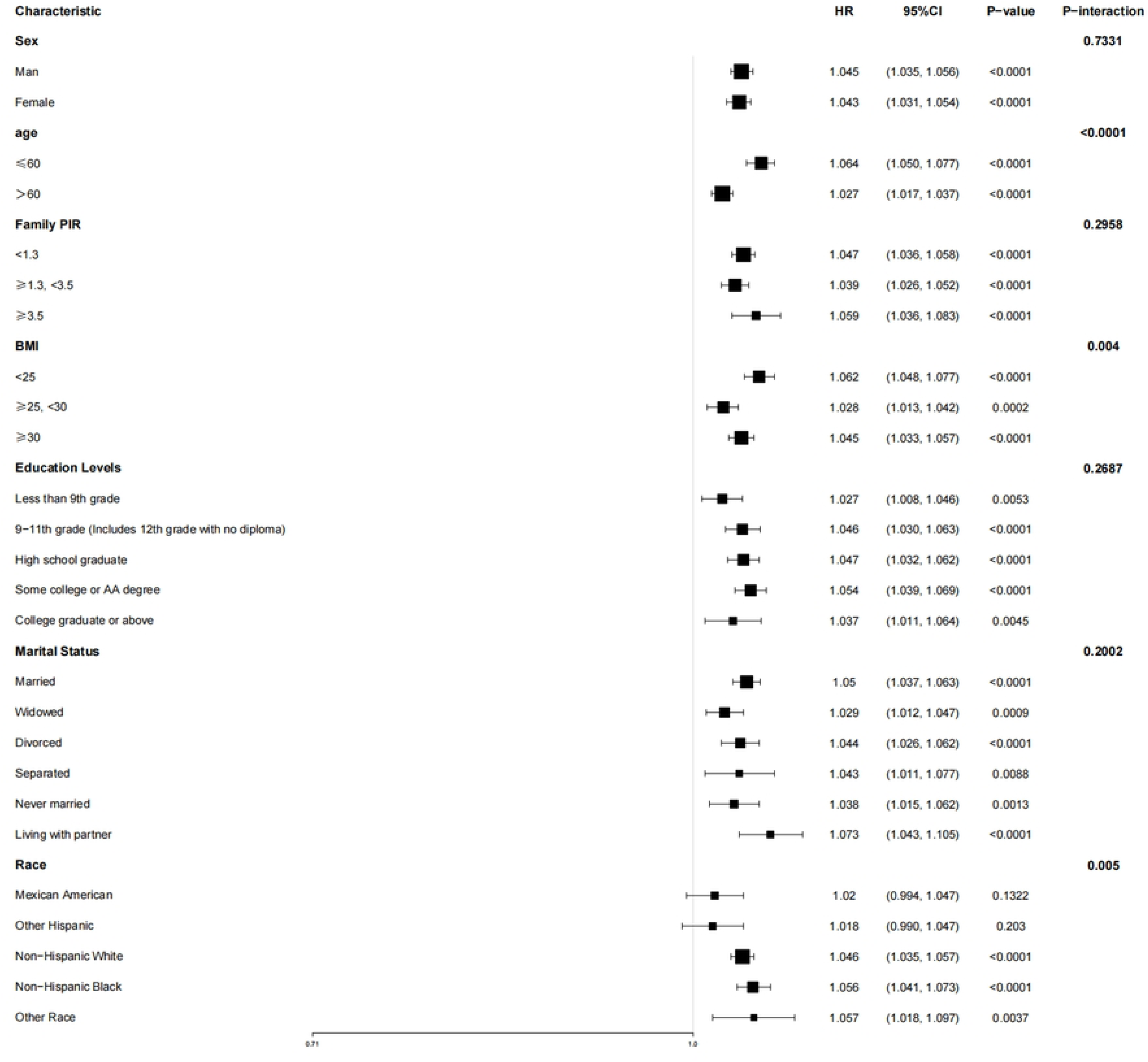
Forest plot of association between depression and all-cause mortality after adjustment for socio-demographic and clinical factors. In addition to the stratification variables themselves, adjustment for sex, age, race, family PIR, education levels, marital status, BMI.

In the supplementary analyses, the trends in the sensitivity analyses were consistent with those in the main analyses. In the supplementary analyses, we used dummy variables to represent missing covariate values. Similar results were obtained after accounting for the effects of missing data (Fig S1 and Table S1).

## Discussion

This study utilised data from the National Health and Nutrition Examination Survey (NHANES) database, spanning from 2005 to 2018, encompassing a total of 36,193 participants. The mean follow-up duration was 90.53 months, during which 3,572 cases of all-cause mortality were documented, constituting a 9.87 all-cause mortality rate. Following adjustment for confounding variables including sex, age, race, family PIR, education levels, marital status, and BMI, the study found that the Patient Health Questionnaire-9 (PHQ-9) depression score exhibited a non-linear relationship with the risk of all-cause mortality, with the mortality risk increasing significantly as the depression score rose. When the PHQ-9 score reached approximately 7 points, the risk growth trend approached saturation. Specifically, when the PHQ-9 score was less than 7 points, the mortality risk increased significantly (HR: 1.068, 95% CI: 1.042–1.095, *P* < 0.0001). Conversely, when the PHQ-9 score exceeded 7 points, the risk increased, albeit to a lesser extent (HR: 1.036, 95% CI: 1.008–1.064, *P* = 0.0113). These findings implied that depressive symptoms are a significant risk factor for all-cause mortality, with a more pronounced risk observed when the PHQ-9 score is below 7 points.The observed associations were consistent across various subgroups and in all conducted sensitivity analyses.

The present study’s findings concerning the overall associations between depression and all-cause mortality risk were consistent with the results of previous studies [24–29]. A number of prospective cohort studies had explored the association between depression and mortality. One such study, based on the NHANES database and including 23,694 participants, demonstrated a significant positive correlation between depression and all-cause mortality [27] .In populations with type 2 diabetes mellitus, depression was associated with an increased risk of all-cause and non-cardiovascular mortality, though no significant association was found with cardiovascular mortality [28]. A further study focusing on asthma patients revealed that depression significantly impacted all-cause mortality risk, particularly worsening the prognosis for asthma patients, suggesting the need for enhanced mental health screening in clinical practice [24]. Furthermore, two large prospective cohort studies – the China Kadoorie Biobank (CKB) and the Dongfeng-Tongji (DFTJ) studies – indicated that depression or depressive symptoms were significantly associated with an increased risk of all-cause mortality, with cardiovascular diseases being the primary direction of association [25]. A multicentre study encompassing 21 countries further demonstrated that individuals exhibiting four or more depressive symptoms had a 20% increased risk of cardiovascular events and death, with this association being more pronounced in urban areas [26].Notably, our study also identified a nonlinear relationship between depression and mortality, determining a threshold value of 7 on the depression score. When depression scores were below 7, the risk growth trend became more significant, potentially due to the advantages of treating depression as a continuous variable through the cox model with restricted cubic spline.

The non-linear relationship between depression and all-cause mortality may be attributed to a confluence of multiple interrelated mechanisms. Foremost among these were biological dysregulation, which can arise from inflammatory processes linked to depression, as evidenced by studies highlighting elevated inflammatory markers in individuals with depressive symptoms, contributing to increased mortality risks [30,31]. Additionally, the cumulative effects of unhealthy behaviors, often exacerbated by depression, such as smoking and poor dietary choices, significantly elevate mortality risk [32]. Incorporating psychological and social factors into this framework underscored how social isolation and lack of support can hinder the management of health conditions effectively [33]. Furthermore, comorbid conditions prevalent among depressed individuals, such as cardiovascular disease and diabetes, not only compound the risk but also accentuate the severity of mortality outcomes, demonstrating a more complex interaction that intensifies at higher levels of depressive severity [34,35]. The notion of biological thresholds becomes particularly relevant here; it suggested that these mechanisms may interact synergistically, culminating in increased mortality risk as the severity of depression intensifies, thereby approaching a saturation point where the additional risk becomes marginally less impactful beyond a certain threshold [36]. Taken together, these findings illustrated a multifaceted and intricate interplay of factors that define the non-linear relationship observed between depression and all-cause mortality.

## Limitations

However, it is imperative to acknowledge the limitations of the study when interpreting the results. Firstly, the depression data were obtained from baseline measurements, which precludes the analysis from explaining the changes in depressive symptoms over time. Secondly, successfully treated participants with depression would not be identified as such by the PHQ-9 criteria. Concurrently, individuals with severe depression may not have participated in the survey or health examination, resulting in an underestimation of the true prevalence of depression. Finally, the study was limited to Americans over the age of 18, so the results may not be generalisable to other populations.

## Conclucion

This study, based on data from the National Health and Nutrition Examination Survey (NHANES), systematically examined the longitudinal association between depressive symptoms and all-cause mortality. The findings revealed a significant nonlinear relationship between depressive symptoms and mortality risk, with evidence of a threshold effect. The increase in mortality risk was more pronounced at lower levels of depressive symptoms, while the risk increment attenuated but remained significant as symptoms worsened.

## Data Availability

All data used in this study are publicly available from the CDC National Center for Health Statistics National Health and Nutrition Examination Survey (NHANES) database (https://www.cdc.gov/nchs/nhanes). Researchers can freely access and download the datasets through the official website.

## Acknowledgements

Not applicable.

## Data availability

The datasets generated and/or analyzed during the current study are publicly available from CDC National Center for Health Statistics https://www.cdc.gov/nchs/nhanes

## Supporting information

**S1 Fig. Association between depression and all-cause mortality**. Adjusted for sex, age, race, family PIR, education level, marital status, BMI.The number of missing values for covariates was as follows: education levels 2,155 (5.95%), marital status 1,634 (4.51%), family PIR 3,061 (8.46%), and BMI 384 (1.06%).The utilisation of dummy variables was necessitated by the absence of continuous variables by more than 1%.

**S1 Table. Threshold effect analysis of depression on the risk of all-cause mortality using two-piecewise regression models, weighted.**Data are presented as HR (95% CI) *P* values; model 1, linear analysis; model 2, nonlinear analysis.Adjusted for sex, age, race ,family PIR, education level, marital status, BMI.The utilisation of dummy variables was necessitated by the absence of continuous variables by more than 1%.The number of missing values for covariates was as follows: education levels 2,155 (5.95%), marital status 1,634 (4.51%), family PIR 3,061 (8.46%), and BMI 384 (1.06%).

